# Prevalence and factors associated with antigen test positivity following SARS-CoV-2 infection among healthcare workers in Los Angeles

**DOI:** 10.1101/2022.07.06.22277341

**Authors:** Paul C. Adamson, Judith S. Currier, Daniel Z. Uslan, Omai B. Garner

## Abstract

Surges of SARS-CoV-2 infections among healthcare workers (HCWs) have led to critical staffing shortages. From January 4 to February 4, 2022, we implemented a return-to-work antigen testing program for HCWs and 870 HCWs participated. Antigen test positivity was 60.5% for those ≤5 days from symptom onset or positive PCR and 47.4% were positive at day 7. Antigen positivity was associated with receiving a booster vaccination and being ≤6 days from symptom onset or PCR test, but not age or a symptomatic infection. Rapid antigen testing can be a useful tool to guide return-to-work and isolation precautions for HCWs following infection.

## Background

In December 2021, a surge of SARS-CoV-2 infections among healthcare workers (HCWs), primarily driven by the Omicron (B.1.1.529) variant, contributed to staffing shortages in health systems across the United States.[1] Following SARS-CoV-2 infection, a period of isolation is recommended to decrease onward viral transmission, the duration of isolation has changed over the course of the COVID-19 pandemic based individual factors including symptoms, vaccination status, and occupation. On December 23, 2021, the U.S. Centers for Disease Control and Prevention modified the isolation guidance for healthcare personnel with SARS-CoV-2 infections, shortening the duration during times of contingency staffing shortages to five days from symptom onset with or without a negative test.[2]

Antigen tests for SARS-CoV-2 detect viral proteins and can provide rapid results. The tests can be performed at the point-of-care or used for at-home testing. Positive antigen tests are associated with higher viral loads and culturable virus, making them useful to identify those more likely to be infectious.[3–7] Thus, antigen tests could be used to discontinue isolation precautions following COVID-19.

The use of antigen testing to guide return-to-work for healthcare workers might be an important tool to reduce the risk for viral transmission to patients and other healthcare workers. This may be especially important during times of high SARS-CoV-2 transmission leading to staffing shortages of healthcare personnel. The aim of this study is to report findings from the implementation of a return-to-work rapid antigen testing program within a large health system, including data on prevalence and factors associated with antigen positivity following COVID-19.

## Methods

On January 4^th^, 2022, University of California, Los Angeles (UCLA) Health implemented an optional, rapid antigen testing program, whereby employees with SARS-CoV-2 infections who were asymptomatic, or mildly symptomatic with improving symptoms, could return to work after 5 days of isolation with a negative antigen test obtained _≥_5 days after their positive PCR test. This is a retrospective analysis of all UCLA Health employees with a positive SARS-CoV-2 PCR test from December 25, 2021 – February 4, 2022 who participated in the return-to-work testing program.

UCLA Health employees access SARS-CoV-2 PCR testing through asymptomatic surveillance testing or based on exposures or self-reported symptoms. Reverse-transcriptase PCR testing was performed by the UCLA Clinical Microbiology Laboratory using the following assays: Simplexa COVID-19 Direct (Diasorin Molecular, Cypress, California), cobas 6800 SARS-CoV-2 and Influenza A/B Test and cobas Liat SARS-CoV-2 and Influenza A/B Assay (Roche Molecular Systems, Pleasanton, California), and TaqMan SARS-CoV-2, FluA/B RT-PCR Assay (ThermoFisher Scientific, Carlsbad, California). Cycle threshold (Ct) values were extracted and analyzed as previously described.[8] The asymptomatic surveillance testing program used SwabSeq, a FDA-authorized high-throughput SARS-CoV-2 testing platform.[9] For the antigen testing program, trained laboratory staff supervised the self-collection of nasal specimens and performed all antigen testing using the Sofia SARS Antigen Fluorescent Immunoassay (Quidel Corporation, Athens, OH).

Employee demographic, vaccination, testing, and symptom data were extracted from electronic employee health records. Symptoms were self-reported using daily symptom surveys. COVID-19 vaccination status at the time of positive PCR test was defined as follows: unvaccinated – if no vaccination was recorded; partially vaccinated – only one vaccine dose was received; fully-vaccinated – primary vaccination series completed > 2 weeks prior; fully vaccinated and boosted – primary vaccination and booster dose was completed > 2 weeks prior.

The primary outcome was antigen positivity by day following symptom onset or positive PCR for those without symptom onset data. Characteristics including age, symptoms, vaccination status, and Ct values were compared between those with positive and negative antigen tests in bivariate and multivariate logistic regression modeling.

The study was approved by the UCLA institutional review board (#21-000373).

## Results

From December 25, 2021 through February 4th, 2022, 2,316 HCWs had a positive SARS-CoV-2 PCR result. Among those, 870 participated in the return-to-work antigen testing program. The median age was 36 years (IQR: 31 – 45). Symptoms were reported by 78.4% (682/870) of those with antigen testing, 9.0% (78/870) reported no symptoms, and symptom data were missing for 12.6% (110/870). COVID-19 vaccination was received by 93.9% (817/870) and 67.4% (586/870) were fully vaccinated and boosted (Table 1).

**Table 1.** SARS-CoV-2 antigen test results and characteristics associated with positivity among 870 healthcare workers participating in a return-to-work antigen testing program in January – February 2022.

| Antigen Test Results |  |  |  |  |  |  |  |  |
| --- | --- | --- | --- | --- | --- | --- | --- | --- |
| (N=870) |  |  |  |  |  |  |  |  |
| Positive/ |  |  |  |  |  |  |  |  |
| Characteristic | Total | (%) | OR | 95% CI | p-value | aOR | 95% CI | p-value |
| Symptomatic |  |  |  |  |  |  |  |  |
| No | 27/74 | 36.5 | Ref | - | - | Ref | - | - |
| Yes | 313/686 | 45.7 | 1.45 | 0.88-2.39 | 0.145 | 2.18 | 0.56, 8.42 | 0.259 |
| Missing | 45/110 | 40.9 | - | - | - | - | - | - |
| Age group, years |  |  |  |  |  |  |  |  |
| 18-44 | 276/659 | 41.9 | Ref | - | - | Ref | - | - |
| 45-60 | 91/183 | 49.7 | 1.35 | 0.97, 1.87 | 0.075 | 1.21 | 0.69, 2.11 | 0.189 |
| ≥ 60 | 18/28 | 64.3 | 2.45 | 1.11, 5.39 | 0.026 | 1.65 | 0.44, 6.16 | 0.456 |
| COVID-19 Vaccination |  |  |  |  |  |  |  |  |
| Status |  |  |  |  |  |  |  |  |
| Unvaccinated | 13/43 | 30.2 | Ref | - | - | Ref | - | - |
| Partially Vaccinated† | 1/1 | 100.0 | - | - | - | - | - | - |
| Fully vaccinated, not |  |  |  |  |  |  |  |  |
| boosted | 105/298 | 35.2 | 1.16 | 0.63, 2.15 | 0.64 | 2.44 | 0.65, 9.23 | 0.189 |
| Fully vaccinated, boosted | 221/418 | 52.9 | 2.35 | 1.28, 4.29 | 0.006 | 4.94 | 1.32, 18.43 | 0.018 |
| Cycle Threshold Values* |  |  |  |  |  |  |  |  |
| <30 | 150/327 | 45.9 | Ref | - | - | Ref | - | - |
| ≥ 30 | 36/69 | 52.2 | 1.28 | 0.76, 2.15 | 0.352 | 1.1 | 0.61, 1.98 | 0.74 |
| Days since onset or test |  |  |  |  |  |  |  |  |
| 5 days or fewer | 159/263 | 60.5 | 7.33 | 4.26,12.61 | <0.001 | 3.70 | 1.68, 8.22 | 0.001 |
| 6 | 85/183 | 46.4 | **4.16** | **2.37, 7.31** | **<0.001** | **2.82** | **1.26, 6.28** | **0.011** |
| 7 | 72/152 | 47.4 | **4.38** | **2.46, 7.82** | **<0.001** | 2.34 | 0.99, 5.50 | 0.053 |
| 8 | 32/93 | 34.4 | **2.62** | **1.37, 5.01** | **0.004** | 1.48 | 0.55, 3.97 | 0.44 |
| 9 | 17/64 | 26.6 | 1.72 | 0.82, 3.58 | 0.149 | 0.82 | 0.28, 2.42 | 0.712 |
| 10 days or more | 20/115 | 17.4 | Ref | Ref | - | Ref | - | - |

†Only 1 observation, logistic regression not performed
\* Cycle threshold values available for Diasorin Simplexa, Roche cobas 6800, and ThermoFisher Taqman assays
Abbreviations: OR - odds ratio; aOR - adjusted odds ratio

Overall, there were 478 (54.9%) negative results, 385 (44.3%) positive results, and 7 (0.8%) indeterminate results. Antigen test positivity following symptom onset or positive PCR test was 61.3% (57/93) for <5 days, 46.5% (308/662) for 5-9 days, and 17.4% (20/115) for _≥_10 days. Figure 1 shows the number of antigen tests performed and the test positivity by day.

**Figure 1.**
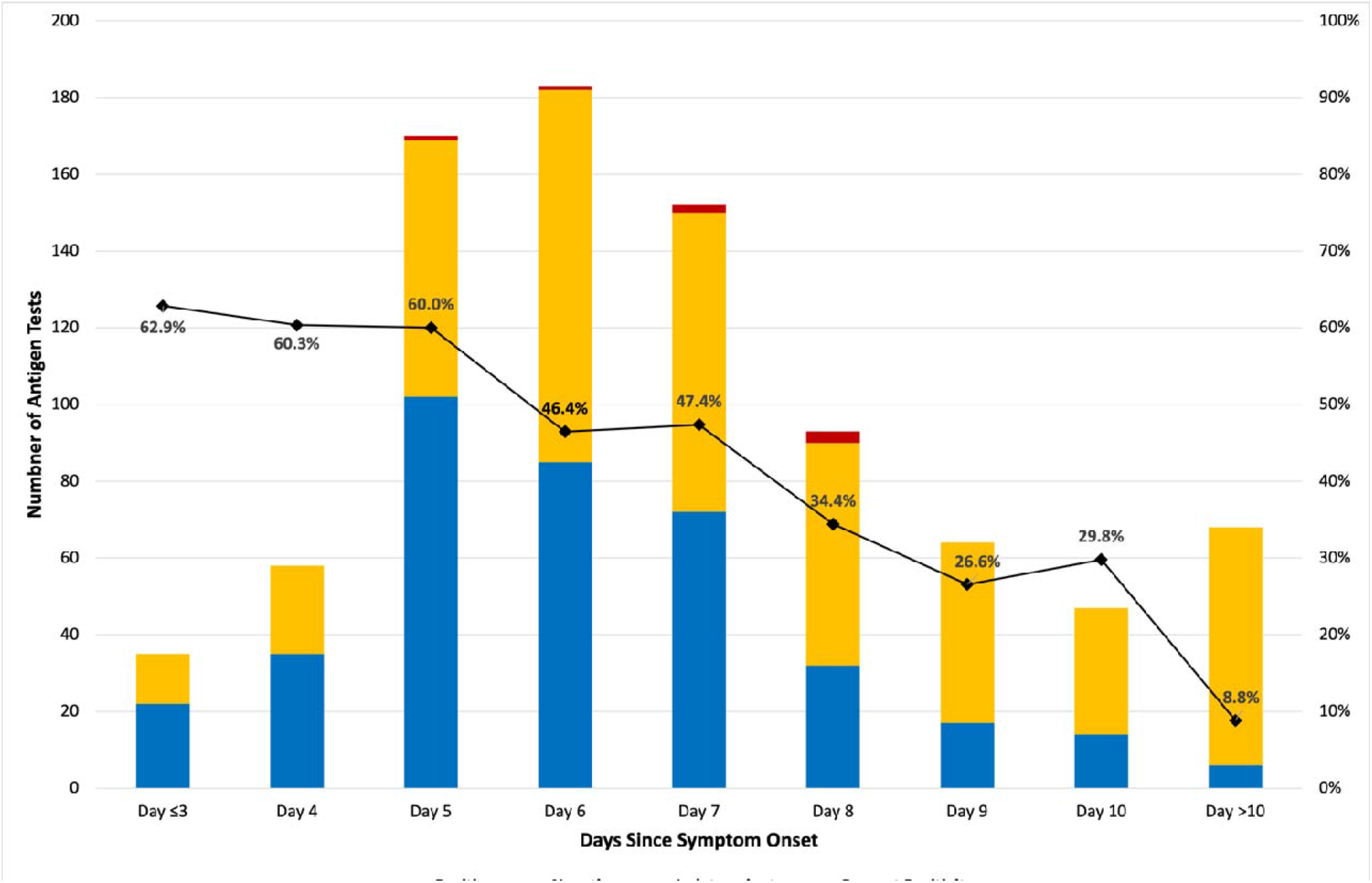
Total number of SARS-CoV-2 antigen tests and positivity by days since symptom onset among healthcare workers.

In multivariate analysis, a positive antigen test was more likely among boosted compared to unvaccinated individuals (adjusted odds ratio [aOR] 4.94; 95% CI: 1.32, 18.43). A positive antigen test was also associated with days following symptom onset or positive PCR test, with those presenting ≤5 days (aOR 3.70; 95% CI: 1.68, 8.2) and on day 6 (aOR 2.82; 95% CI: 1.26, 6.28) being more likely to have a positive test. (Table 1)

## Discussion

In this retrospective, observational study during a time when nearly all SARS-CoV-2 infections were the Omicron variant,[10] 60% of asymptomatic or mildly symptomatic HCWs participating in the return-to-work antigen testing program were positive 5 days following symptom onset or positive PCR and 50% were positive on day 7. Being fully vaccinated and boosted was associated with increased odds of a positive antigen test. Testing ≤6 days following symptom onset or a positive PCR was also associated with increased odds of a positive antigen test. Antigen test positivity was not associated with Ct values <30 or with symptomatic infections.

We found a high proportion of antigen positivity among HCWs on days 5-7 following onset of symptoms or a positive PCR. Data on the use of rapid antigen testing among healthcare workers following COVID-19 are limited and estimates vary. In one study among 290 HCWs, 49% of those testing on Day 5 after COVID-19 diagnosis were antigen positive and there was no difference among those unvaccinated compared to fully vaccinated and/or boosted.[11] In a larger sample of 1661 HCWs, 88.1% were antigen positive on Day 5 and antigen positivity was associated with symptomatic infections and vaccinations being up to date.[12] Our findings on antigen-positivity falls between these two prior reports and show that a majority of HCWs might continue to be infectious at 5 days.

Similar to Tande et al., we found that being fully vaccinated and boosted was associated with antigen positivity compared to being unvaccinated.[12] While data demonstrate that boosted individuals were less likely to be infected with Omicron,[13] emerging data suggest those with breakthrough infections might have delayed viral decay and a longer time to viral clearance, which might explain our findings.[14, 15] However, we did not perform viral culture or repeat PCR testing as part of this study. Since this was an optional testing program, it is also possible that unvaccinated HCWs were more likely to have symptomatic infections that made them ineligible to participate in return-to-work testing, so findings should be interpreted within this context.

This report on antigen test positivity among HCWs participating in a return-to-work testing program within a large health system provides important data for healthcare settings. Antigen testing prior to shortening the duration of isolation can risk-stratify HCWs before returning to work, allowing for interventions to mitigate viral transmission in healthcare settings, including the use of high-filtration masks, repeat antigen testing, or extending the duration of isolation.

## Data Availability

All data produced in the present study are available upon reasonable request to the authors.

